# Epigenetic Signatures in Monozygotic and Dizygotic Twins Discordant for Orofacial Clefts

**DOI:** 10.64898/2026.04.07.26350251

**Authors:** A. L. Petrin, H. L. Keen, L. Dunlay, X. J. Xie, E. Zeng, A. Butali, A. Wilcox, M. L. Marazita, J. C. Murray, L. M. Moreno-Uribe

## Abstract

**Introduction:** Nonsyndromic cleft lip with or without cleft palate (NSCL/P) is a common congenital malformation with complex etiology involving both genetic and environmental factors. Epigenetic mechanisms may mediate environmental contributions, but separating genetic from environmental effects remains challenging.

**Methods:** We present an epigenome-wide association study with 32 monozygotic and 22 dizygotic twin pairs discordant for NSCL/P on blood and saliva samples. Differential methylation analysis was conducted using linear models to identify CpG sites showing significant methylation differences between affected and unaffected twins followed by functional annotation and pathway enrichment analysis.

**Results:** The top-ranked finding is a differentially methylated region comprising two CpG sites at the *CYP26A1* locus, cg12110262 (P = 3.21x10^-7^) and cg15055355 (P = 1.39x10^-3^). *CYP26A1* is essential for retinoic acid catabolism and craniofacial patterning. The chromatin regulator *ANKRD11*, which causes KBG syndrome featuring cleft palate was the second best hit. Differentially methylated CpG sites showed significant enrichment in craniofacial enhancers and overlap with multiple GWAS-validated cleft genes including *VAX1, PVRL1, SMAD3*, and *PRDM16*.

**Conclusions:** Our findings implicate retinoic acid signaling and chromatin regulation in NSCL/P etiology and demonstrate the value of discordant twin designs for distinguishing environmental from genetic epigenetic contributions to complex malformations.

## Introduction

Nonsyndromic cleft lip with or without cleft palate (NSCL/P) represents one of the most prevalent congenital malformations worldwide, affecting approximately 1 in 700 live births with significant variation across ethnic populations^1^. Despite decades of research, the complex etiology of NSCL/P remains incompletely understood, involving intricate interactions between genetic susceptibility, environmental exposures, and epigenetic modifications^2,3^. While genome-wide association studies have identified numerous genetic risk loci explaining approximately 25% of NSCL/P heritability^4,5^, the substantial “missing heritability” suggests that epigenetic mechanisms may play a critical role in disease pathogenesis.

DNA methylation, the most extensively studied epigenetic modification, serves as a dynamic interface between genetic predisposition and environmental influences during embryonic development^6^. Palatogenesis requires precise spatiotemporal coordination of cellular processes including neural crest migration, proliferation, differentiation, and apoptosis—all of which are subject to epigenetic regulation^7,8^. Environmental factors known to influence NSCL/P risk, including maternal smoking, alcohol consumption, and nutritional deficiencies, can induce methylation changes that persist throughout development^9,10^.

Previous methylation studies in NSCL/P have identified differential methylation in genes encoding transcription factors (*LHX8, PRDM16, PBX1*), growth factors (*WNT9B*, *BMP4*), and microRNAs (*MIR140*, *MIR300*)^11,12^. However, these studies have been limited by small sample sizes, candidate gene approaches, or inability to control for genetic background effects.

In a landmark study of monozygotic twins discordant for Van der Woude syndrome, we identified significant DNA methylation differences at CpG sites near *TP63*, which functions in a regulatory loop with *IRF6* to coordinate epithelial proliferation and differentiation during palatogenesis^13^. Another study on a few pairs of twins discordant for NSCL/P using whole genome bisulfite sequencing revealed differentially methylated regions in candidate genes including *MAFB* and *ZEB2*, as well as differential methylation in genes belonging to the Hippo signaling pathway^14^.

In this study, we used the Illumina EPIC array to analyze methylation patterns in blood and saliva samples from both monozygotic and dizygotic twin pairs, employing rigorous statistical approaches to identify disease-associated epigenetic variation. Tissue selection represents a critical consideration in methylation studies of developmental disorders and the comparative analysis of blood and saliva methylation patterns may reveal tissue-specific epigenetic mechanisms while informing the utility of different sample types for biomarker development.

The discordant twin design offers unique advantages for epigenetic studies of complex traits ^15,16^. Monozygotic (MZ) twins share essentially identical genomes, so methylation differences between affected and unaffected co-twins must arise from environmental, stochastic, or developmental mechanisms rather than genetic variation. Dizygotic (DZ) twins share approximately 50% of their genetic variation, allowing assessment of how genetic background influences epigenetic susceptibility. Comparing MZ and DZ findings enables separation of purely environmental epigenetic effects from gene-environment interactions, and may reveal variance methylation quantitative trait loci (VmeQTLs)—genetic variants that modulate epigenetic plasticity rather than mean methylation levels^17,18^.

Our study addresses several critical gaps in the understanding of NSCL/P etiology through comprehensive epigenome analysis of twins discordant for cleft lip and palate. We profiled DNA methylation in blood and saliva samples from 32 MZ and 22 DZ twin pairs using the Illumina MethylationEPIC array. Our analysis identifies differentially methylated CpG sites implicating retinoic acid signaling and chromatin regulatory mechanisms in cleft etiology, validates findings through known cleft susceptibility genes, and reveals distinct epigenetic patterns between MZ and DZ twins suggesting multiple epigenetic paths to this common malformation.

## Results

### Study population and methylation profiling

We enrolled 32 pairs of MZ twins and 22 pairs of DZ twins discordant for NSCL/P and . Zygosity was confirmed using standard genetic markers. The cohort included 58 males and 50 females, with cleft phenotypes comprising cleft lip and palate (CLP, n=37) and cleft lip only (CL, n=17). Blood samples were available for 24 MZ pairs and 9 DZ pairs, while saliva samples were collected from 8 MZ pairs and 13 DZ pairs.

Genome-wide DNA methylation was assessed using the Illumina Infinium MethylationEPIC BeadChip array, interrogating over 850,000 CpG sites with enhanced coverage of enhancer regions and regulatory elements compared to previous arrays. After quality control, normalization, and removal of probes with known SNPs at CpG sites, differential methylation analysis was conducted using limma linear models accounting for the paired nature of twin samples. Analyses were stratified by zygosity (MZ vs. DZ) and tissue type (blood vs. saliva).

### Overview of differential methylation

We identified substantial numbers of differentially methylated CpG sites (DMCs) across all four analyses (Fig. 1 and 2, Table 1). In MZ blood, 2,578 DMCs reached nominal significance (P < 0.05), with 845 at P < 0.001 and 114 at P < 0.0001. DZ blood analysis identified 1,826 DMCs (370 at P < 0.001, 35 at P < 0.0001). MZ saliva yielded the largest number of DMCs (n = 3,254; 1,124 at P < 0.001), while DZ saliva identified 1,783 DMCs (505 at P < 0.001).

**Figure 1:**
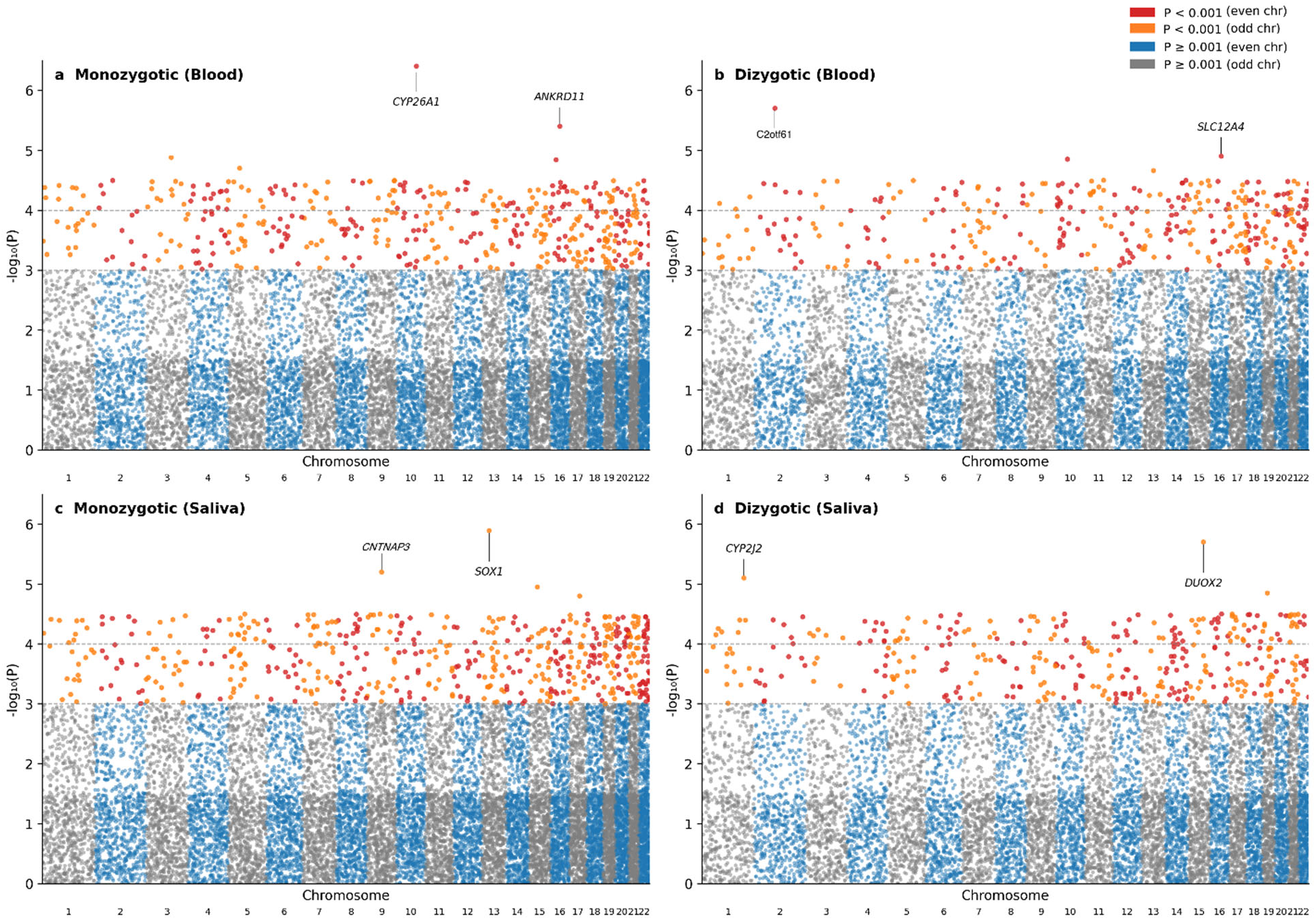
Manhattan plots of epigenome-wide association results. Manhattan plots displaying -log10(P-value) for each CpG site across chromosomes 1-22 for (a) monozygotic twins blood (n=24 pairs), (b) dizygotic twins blood (n=9 pairs), (c) monozygotic twins saliva (n=8 pairs), and (d) dizygotic twins saliva (n=13 pairs) analyses. Horizontal dashed lines indicate significance thresholds at P = 1 × 10-3 (upper) and P = 1 × 10-4 (lower). Top hits are labeled with nearest gene names based on GREAT annotation.

**Figure 2:**
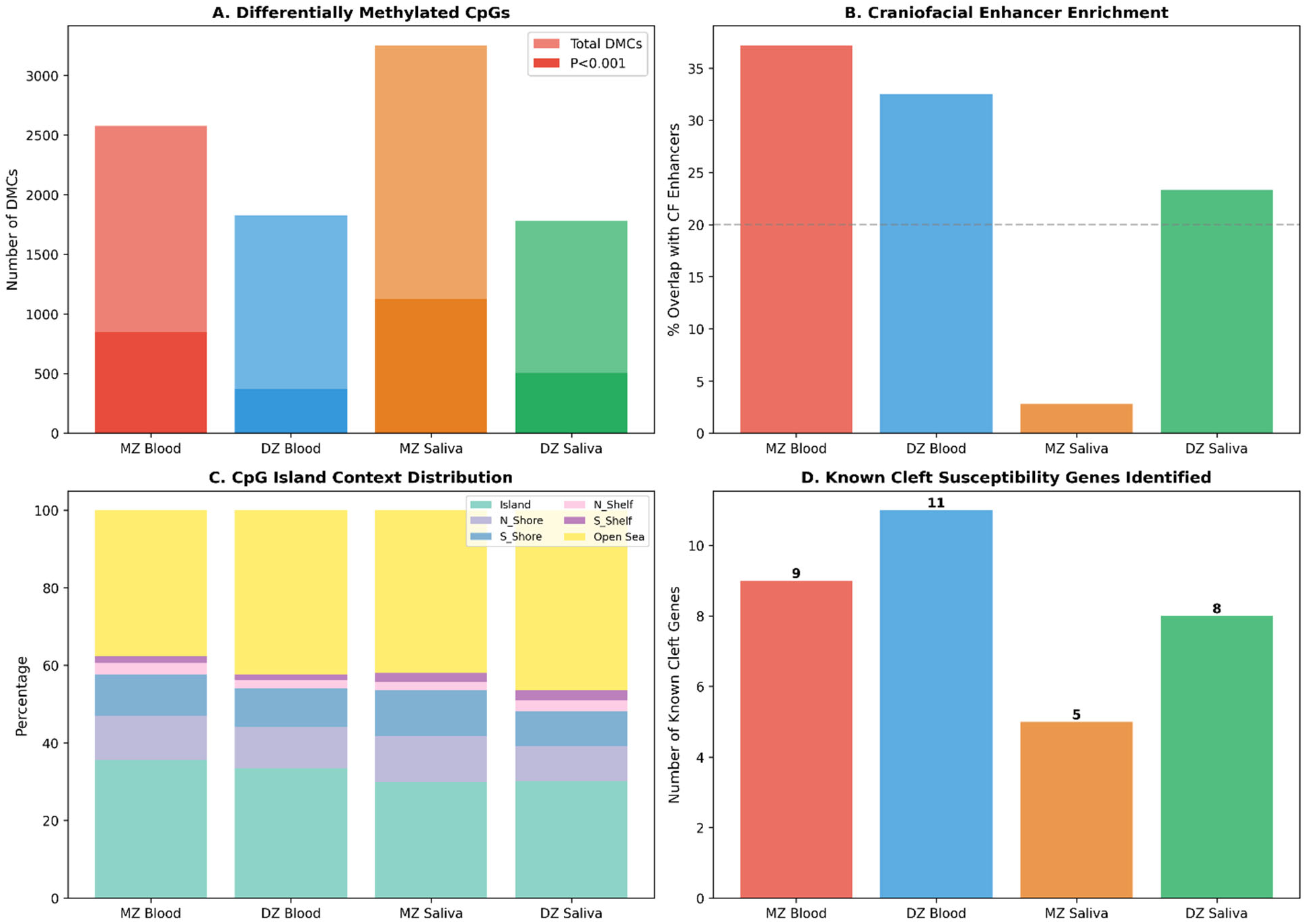
Comparative analysis of differential methylation findings across zygosity and tissue types. (A) Differentially methylated CpG sites (DMCs) identified in each analysis. Bar plot showing the total number of DMCs at nominal significance (P < 0.05, light blue) and stringent threshold (P < 0.001, dark blue) for each analysis. (B) Enrichment of DMCs in craniofacial enhancer elements. Percentage of DMCs (P < 0.05) overlapping with annotated craniofacial enhancers derived from developmental chromatin profiling studies. Dashed line indicates the genomic background rate (∼20%). (C) Distribution of DMCs across CpG island context. Stacked bar plot showing the proportion of DMCs (P < 0.001) mapping to CpG islands, shores (regions flanking islands, N_Shore and S_Shore), shelves (regions flanking shores, N_Shelf and S_Shelf), and open sea (isolated CpGs). (D) Identification of known cleft susceptibility genes. Number of established cleft susceptibility genes (from a curated list of 48 genes identified through GWAS, linkage studies, and syndromic cleft gene discovery) harboring at least one DMC (P < 0.05) in each analysis.

**Table 1:** Twin pairs by tissue type and number of DMCs across all groups.

| Analysis | Twin Pairs | Total DMCs<br>($P < 0.05$) | DMCs<br>($P < 0.001$) | DMCs<br>($P < 0.0001$) | DMCs<br>($P < 0.00001$) | CF Enhancer<br>Overlap (%) | Known Cleft<br>Genes | CpG Islands<br>(%) |
| --- | --- | --- | --- | --- | --- | --- | --- | --- |
| MZ Blood | 24 | 2578 | 845 | 114 | 14 | 37.2% | 9 | 35.6% |
| DZ Blood | 9 | 1826 | 370 | 35 | 4 | 32.5% | 11 | 33.4% |
| MZ Saliva | 8 | 3254 | 1124 | 101 | 10 | 2.8% | 5 | 29.8% |
| DZ Saliva | 13 | 1783 | 505 | 44 | 5 | 23.3% | 8 | 30.2% |

MZ; monozygotic; DZ: dizygotic; DMC: differentially methylated CpG sites; CF: craniofacial

The genomic distribution of top DMCs showed characteristic patterns across analyses (Table 1 and Fig. 2c). Approximately 30-36% of DMCs at P < 0.001 were located within CpG islands, consistent with involvement of gene promoters and transcription start sites where methylation typically exerts direct transcriptional effects. An additional 15-20% of DMCs mapped to CpG island shores (N_Shore and S_Shore), regions flanking CpG islands that are known for tissue-specific methylation variation and have been implicated in developmental gene regulation.

Notably, 40-50% of top DMCs were located in open sea regions—isolated CpGs far from CpG islands. Many of these open sea DMCs overlapped with annotated enhancer elements, highlighting the importance of distal regulatory regions in cleft-associated epigenetic variation. A smaller proportion (5-8%) mapped to CpG island shelves. This distribution, with substantial representation across promoter-associated islands, tissue-variable shores, and enhancer-rich open sea regions, suggests that cleft-associated methylation differences affect multiple layers of gene regulation.

Gene region analysis showed enrichment in gene bodies (40-50% of DMCs) and promoter regions (TSS1500, TSS200; 15-25%), with additional representation in 5’UTR/first exon regions (10-15%). The predominance of gene body methylation is consistent with established patterns in EWAS, where intragenic methylation often correlates positively with gene expression and may reflect transcriptional elongation or alternative promoter usage.

### Top differentially methylated CpG sites in monozygotic twins

The most significant finding in MZ blood was a differentially methylated region (DMR) at the *CYP26A1* locus on chromosome 10, comprising two CpG sites with distinct regulatory contexts (Fig. 1). The lead CpG, cg12110262 (P = 3.21 x 10^-7^; position 94,827,200), lies within an annotated craniofacial enhancer element in the intergenic region between *CYP26A1* and *CYP26C1*. The second CpG, cg15055355 (P = 1.39 x 10^-3^; position 94,833,626), is located just 21 bp upstream of the *CYP26A1* transcription start site (TSS), directly within the core promoter region. These two CpG sites span approximately 6.4 kb and together implicate coordinated epigenetic dysregulation affecting both a distal enhancer element and the proximal promoter of *CYP26A1*.

*CYP26A1* encodes cytochrome P450 family 26 subfamily A member 1, a critical enzyme for retinoic acid (RA) catabolism. RA signaling is essential for craniofacial patterning during embryonic development, establishing anteroposterior gradients required for proper facial morphogenesis. *CYP26* enzymes degrade retinoic acid to create RA-free zones necessary for tissue specification, and disruption of *CYP26A1* causes craniofacial malformations including cleft palate in animal models ^19,20^. The identification of differential methylation at both an enhancer element and the core promoter of *CYP26A1* in genetically identical MZ twins provides strong evidence for environmental or stochastic epigenetic perturbation of this critical developmental pathway. The presence of two independent CpG sites at this locus substantially strengthens confidence in this finding and suggests a mechanism whereby methylation changes could directly impact *CYP26A1* transcriptional regulation.

The second-ranked hit in MZ blood, cg08392591 (P = 4.01 x 10^-6^), maps directly to *ANKRD11*, a known cleft susceptibility gene. Mutations in *ANKRD11* cause KBG syndrome (OMIM #148050), characterized by macrodontia, distinctive facial features, and in approximately 30% of cases, cleft palate ^21^. *ANKRD11* encodes a chromatin regulator that modulates histone acetylation at gene enhancers through interaction with histone deacetylases and p160 coactivators ^22^. Additional top MZ blood hits included *PIK3CA* (P = 1.33 x 10^-5^), a *PI3K* signaling component with known roles in craniofacial overgrowth syndromes.

In MZ saliva, the top hit (cg09469566, P = 1.29 x 10^-6^) mapped near *SOX1*, a *SRY*-related *HMG*-box transcription factor involved in neural development. Notable findings included DMCs associated with *LHX8* (P = 2.03 x 10^-4^), a LIM homeobox gene essential for orofacial (including palate) and tooth development ^23^; *WT1* (P = 1.86 x 10^-4^), associated with WAGR syndrome (which presents craniofacial features); and *GPC3* (P = 2.47 x 10^-4^), encoding glypican-3, mutations of which cause Simpson-Golabi-Behmel syndrome featuring characteristic facial features. The top ten hits for each group are summarized in Table 2.

**Table 2.**
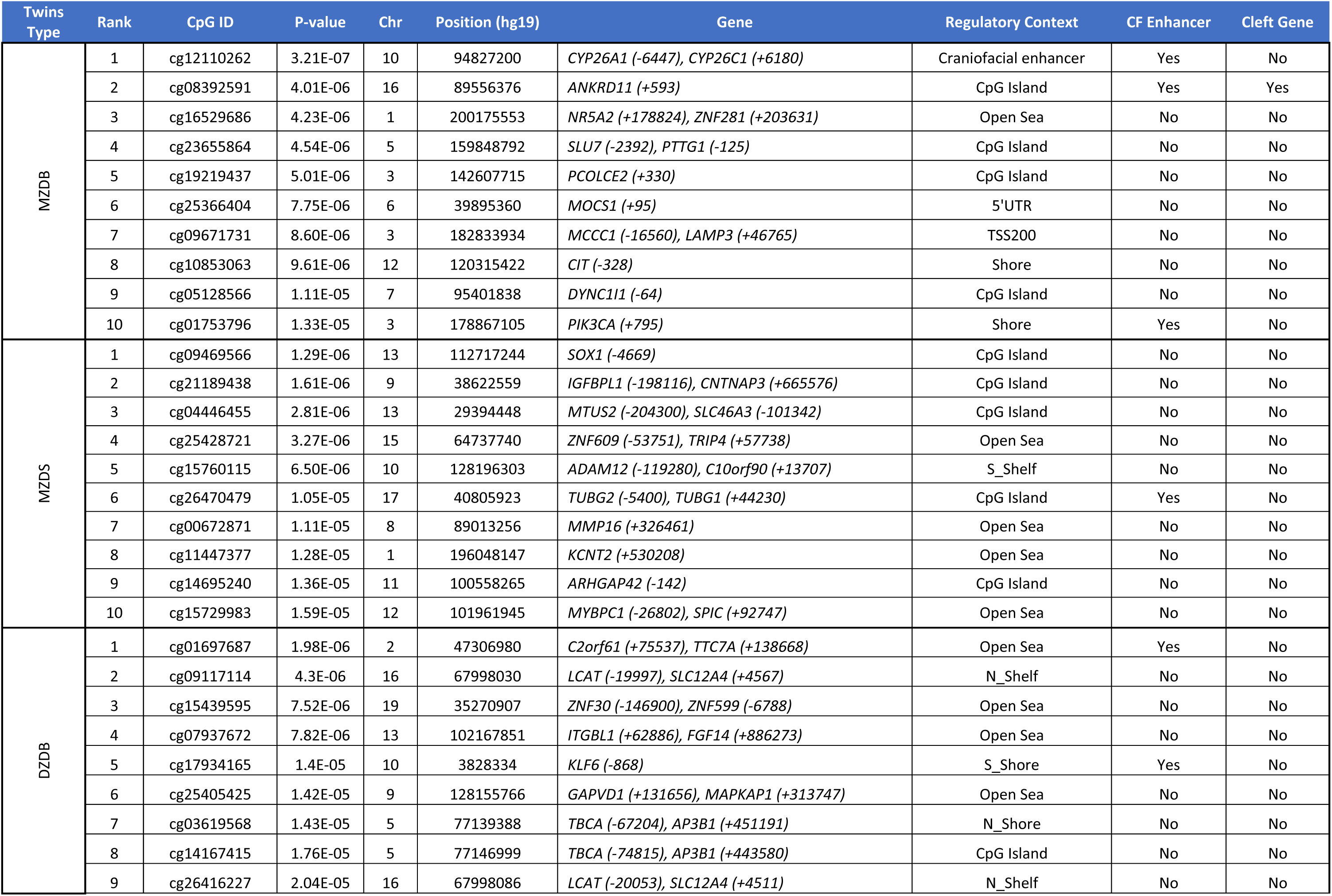
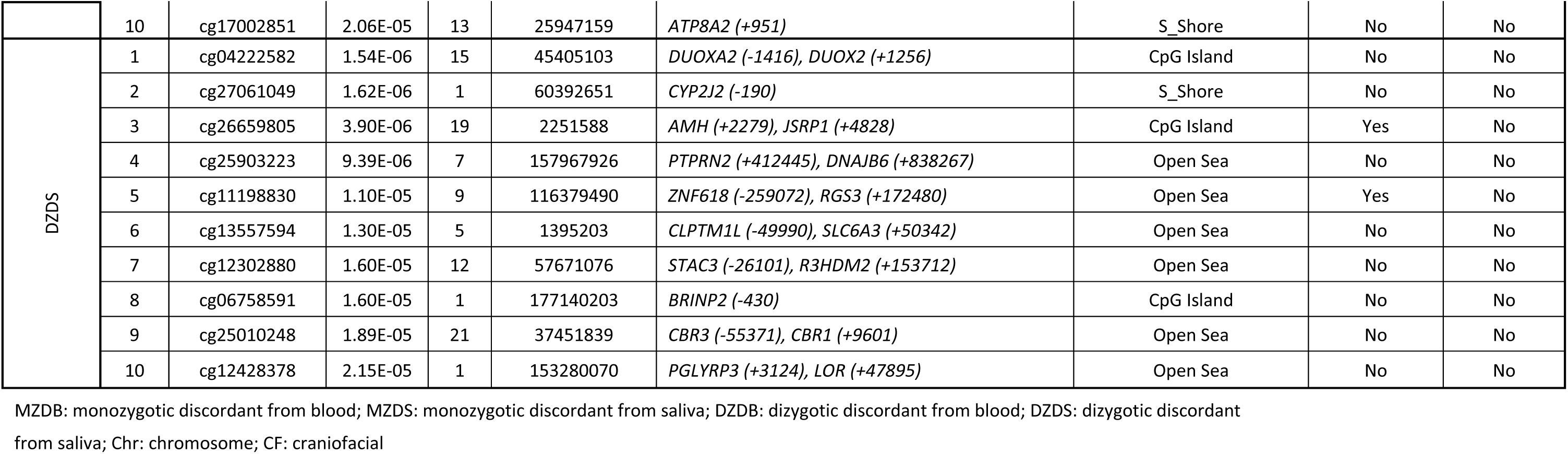
Top 10 CpG sites per twin group.

### Top differentially methylated CpG sites in dizygotic twins

The DZ blood analysis identified distinct genomic associations. The top hit (cg01697687, P = 1.98 x 10^-6^) mapped to an intergenic region within an annotated craniofacial enhancer. Notable gene-associated findings included *KLF6* (P = 1.40 x 10^-5^), a Kruppel-like transcription factor involved in cell differentiation, and *TNXB* (P = 2.20 x 10^-5^), encoding tenascin-XB, an extracellular matrix protein. DZ blood analysis identified several known cleft genes including *PITX2* (P = 1.36 x 10^-3^) and *PRDM16* (P = 7.49 x 10^-4^). DZ saliva analysis yielded notable associations with established cleft susceptibility genes. *VAX1* (P = 6.42 x 10^-4^), encoding ventral anterior homeobox 1, is a GWAS-validated cleft susceptibility gene essential for facial patterning^5^. *PVRL1/NECTIN1* (P = 4.48 x 10^-4^) encodes nectin-1, mutations of which cause *CLPED1* syndrome featuring cleft lip/palate ^24^. *CDK6* (P = 1.04 x 10^-4^), a cell cycle regulator, has been associated with Van der Woude syndrome modifier effects. *ALX4* (P = 9.32 x 10^-4^), encoding aristaless-like homeobox 4, is mutated in frontonasal dysplasia with craniofacial involvement. As mentioned above, the top ten hits for each group are summarized in Table 2.

### Comparison of monozygotic and dizygotic twin findings

A striking observation was the minimal overlap between MZ and DZ analyses (Fig. 3). At P < 0.001, only 3 CpGs were shared between MZ blood and DZ blood analyses, and only 2 CpGs overlapped between MZ saliva and DZ saliva. Within the same zygosity, 3 CpGs overlapped between MZ blood and MZ saliva, while no CpGs overlapped between DZ blood and DZ saliva. Overall, of the 2,834 unique DMCs at P < 0.001 across all analyses, only 7 were identified in more than one analysis (MZ-specific: 1,959 CpGs; DZ-specific: 868 CpGs).

**Figure 3:**
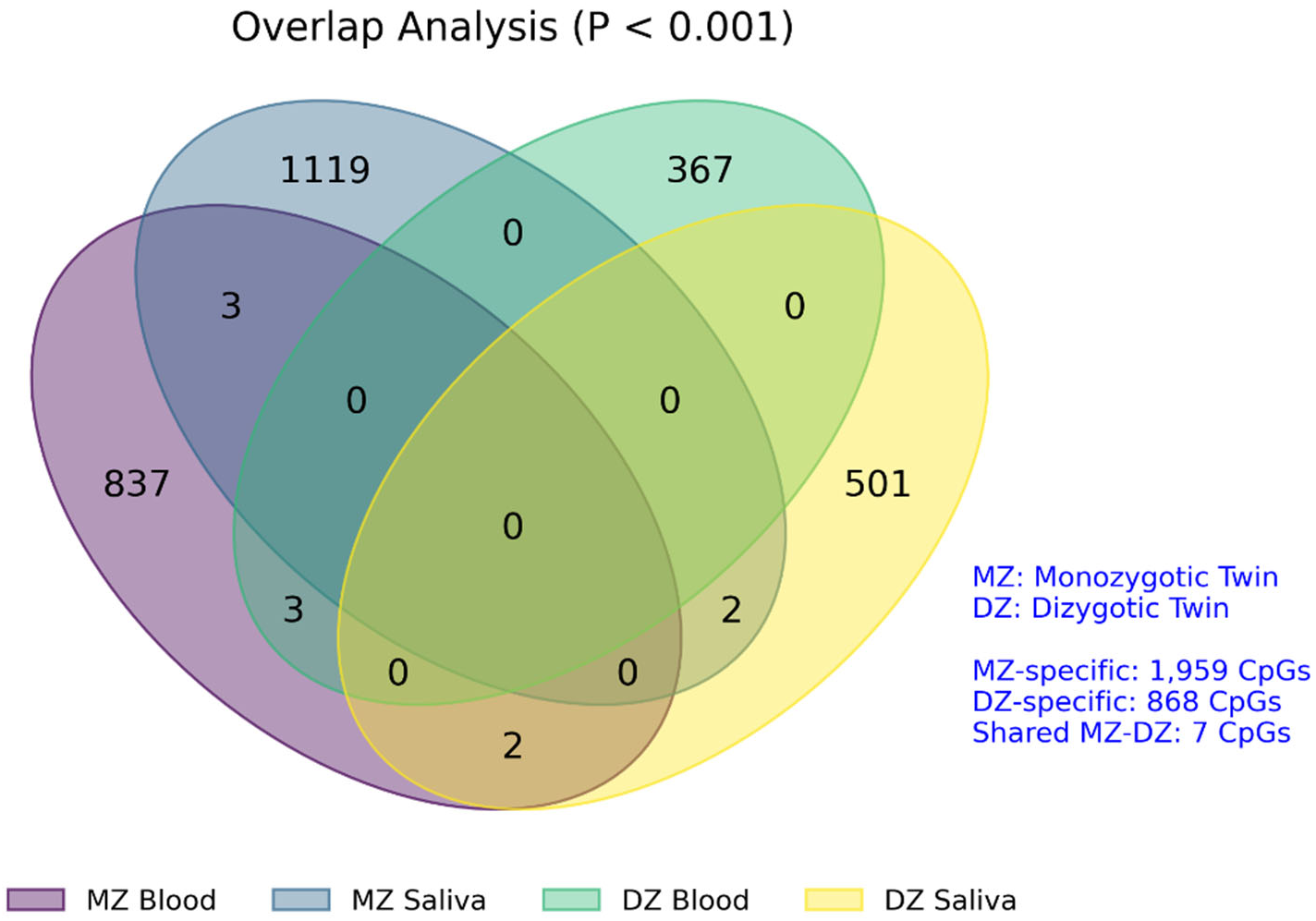
Overlap analysis reveals distinct epigenetic signatures between twin types. Venn diagram illustrating the overlap of differentially methylated CpG sites (DMCs) at P < 0.001 across the four analyses.

Despite limited CpG-level overlap, both MZ and DZ analyses consistently identified genes from known cleft-associated pathways, suggesting convergence at the biological pathway level rather than individual CpG sites. MZ analyses identified 12 cleft-associated genes in blood and 11 in saliva, while DZ analyses identified 6 genes in blood and 9 in saliva, with *PRDM16* identified across multiple analyses.

### Enrichment in craniofacial regulatory elements

We assessed overlap of top DMCs with annotated craniofacial enhancers derived from developmental chromatin profiling studies (Fig. 2b). MZ blood showed striking enrichment, with 37.2% of DMCs (958/2,578) overlapping craniofacial enhancer elements, substantially exceeding the background rate of approximately 20%. DZ blood showed similar enrichment at 32.5% (594/1,826).

Interestingly, MZ saliva showed minimal enhancer overlap (2.8%, 92/3,254), while DZ saliva showed intermediate enrichment (23.3%, 415/1,783).

The higher craniofacial enhancer enrichment in blood compared to saliva, particularly for MZ twins, suggests that systemic methylation patterns in blood may more directly reflect developmental programming at tissue-specific regulatory elements. The *CYP26A1* DMR exemplifies this pattern, with the lead CpG (cg12110262) lying within a craniofacial enhancer and the second CpG (cg15055355) positioned at the core promoter, together suggesting coordinated epigenetic control of this critical developmental gene.

### Identification of known cleft susceptibility genes

To validate our findings, we examined overlap with known cleft-associated genes identified through GWAS, linkage studies, and syndromic cleft gene discovery (Fig. 2d, Table 2). Across all analyses, 25 known cleft genes harbored at least one DMC, providing strong biological validation of the study approach. Notable findings included multiple DMCs at *ANKRD11* (3 CpGs in MZ blood, P^top^ = 4.01 x 10^-6^; 2 CpGs in MZ saliva), *PRDM16* (present in all four analyses), and *PVRL1* (MZ saliva and DZ saliva).

Several genes showed analysis-specific associations: *SMAD3*, a *TGF*-beta signaling component essential for palatal fusion, was identified only in MZ blood (P = 2.03 x 10^-4^). *VAX1*, the GWAS-validated homeobox gene, was specific to DZ saliva (P = 6.42 x 10^-4^). *IRF6*, the most replicated cleft susceptibility gene, showed nominal association in DZ blood (P = 4.53 x 10^-3^) and MZ saliva (P = 1.31 x 10^-3^). These patterns suggest that different genetic backgrounds (MZ vs. DZ) and tissue contexts (blood vs. saliva) capture distinct aspects of cleft-associated epigenetic variation. Table 3 shows the 25 cleft-associated genes that harbored at least one DMC (p<0.05) across all analyses.

**Table 3:** 25 known cleft susceptibility genes harboring at least one DMC (P<0.05) across all analyses.

| Gene | MZ Blood CpGs | MZ Blood Top P | DZ Blood CpGs | DZ Blood Top P | MZ Saliva CpGs | MZ Saliva Top P | DZ Saliva CpGs | DZ Saliva Top P | Biological Function |
| --- | --- | --- | --- | --- | --- | --- | --- | --- | --- |
| *ANKRD11* | 3 | $4.01 \times 10^{-6}$ | - | - | 2 | $7.78 \times 10^{-4}$ | - | - | Chromatin regulation; KBG syndrome |
| *PRDM16* | 3 | $3.27 \times 10^{-5}$ | 3 | $7.49 \times 10^{-4}$ | 2 | $9.94 \times 10^{-4}$ | 1 | $2.32 \times 10^{-3}$ | Transcriptional regulation; GWAS locus |
| *DLX6* | 2 | $3.69 \times 10^{-4}$ | - | - | - | - | - | - | Homeobox TF; craniofacial development |
| *MLL2/KMT2D* | 1 | $3.41 \times 10^{-4}$ | - | - | - | - | - | - | H3K4 methyltransferase; Kabuki syndrome |
| *SMAD3* | 1 | $2.03 \times 10^{-4}$ | - | - | - | - | - | - | TGF-β signaling; palatal fusion |
| *TFAP2A* | 1 | $1.03 \times 10^{-3}$ | 1 | $8.27 \times 10^{-3}$ | - | - | - | - | AP-2α; BOFS syndrome |
| *MSX2* | 1 | $1.17 \times 10^{-3}$ | - | - | - | - | - | - | Homeobox TF; craniosynostosis |
| *PITX2* | 1 | $2.35 \times 10^{-3}$ | 1 | $1.36 \times 10^{-3}$ | - | - | - | - | Homeobox TF; Rieger syndrome |
| *FGFR1* | 1 | $1.62 \times 10^{-3}$ | - | - | - | - | - | - | FGF receptor; Kallmann/Pfeiffer syndrome |
| *PAX9* | 1 | $1.86 \times 10^{-3}$ | - | - | 1 | $7.27 \times 10^{-4}$ | - | - | Paired box TF; tooth agenesis |
| *ALX3* | 1 | $3.36 \times 10^{-3}$ | - | - | - | - | - | - | Homeobox TF; frontonasal dysplasia |
| *SPECC1L* | 1 | $3.76 \times 10^{-3}$ | - | - | 1 | $2.21 \times 10^{-4}$ | - | - | Cytoskeleton; oblique facial clefts |
| *IRF6* | - | - | 1 | $4.53 \times 10^{-3}$ | 1 | $1.31 \times 10^{-3}$ | - | - | Transcription factor; VWS/PPS |
| *BMP4* | - | - | 1 | $5.55 \times 10^{-3}$ | - | - | - | - | BMP signaling; orofacial clefting |
| *WNT3* | - | - | 1 | $6.91 \times 10^{-3}$ | 2 | $5.48 \times 10^{-4}$ | 1 | $6.88 \times 10^{-4}$ | Wnt signaling; tetra-amelia |
| *LHX8* | - | - | - | - | 1 | $2.03 \times 10^{-4}$ | 1 | $1.35 \times 10^{-3}$ | LIM homeobox; tooth development |
| *SOX9* | - | - | - | - | 1 | $8.74 \times 10^{-4}$ | - | - | HMG-box TF; campomelic dysplasia |
| *CTNND1* | - | - | - | - | 1 | $1.22 \times 10^{-3}$ | - | - | Catenin delta-1; blepharocheilodontic syndrome |
| *PVRL1* | - | - | - | - | 1 | $2.24 \times 10^{-3}$ | 1 | $4.48 \times 10^{-4}$ | Cell adhesion; CLPED1 syndrome |
| *MAFB* | - | - | - | - | 1 | $2.42 \times 10^{-3}$ | - | - | Multicentric carpotarsal osteolysis |
| *CDK6* | - | - | - | - | - | - | 1 | $1.04 \times 10^{-4}$ | Cell cycle; VWS modifier |
| *VAX1* | - | - | - | - | - | - | 1 | $6.42 \times 10^{-4}$ | Homeobox TF; GWAS locus |
| *ALX4* | - | - | - | - | - | - | 1 | $9.32 \times 10^{-4}$ | Homeobox TF; frontonasal dysplasia |
| *MSX1* | - | - | - | - | - | - | 1 | $1.71 \times 10^{-3}$ | Homeobox TF; tooth agenesis/clefting |
| *SATB2* | - | - | - | - | - | - | 1 | $3.64 \times 10^{-3}$ | Chromatin organizer; Glass syndrome |

MZ: monozygotic; DZ: Dizygotic

### Pathway enrichment analysis

Gene set enrichment analysis using genes annotated to top DMCs revealed enrichment in biologically relevant pathways (Fig. 4). MZ blood showed significant enrichment for known cleft susceptibility genes (fold enrichment = 5.69, P = 4.76 x 10^-2^, hypergeometric test), with *ANKRD11* and *PRDM16* driving this signal. DZ saliva showed stronger enrichment (fold enrichment = 9.27, P = 3.96 x 10^-3^) driven by *VAX1, PVRL1*, and *CDK6*.

**Figure 4.**
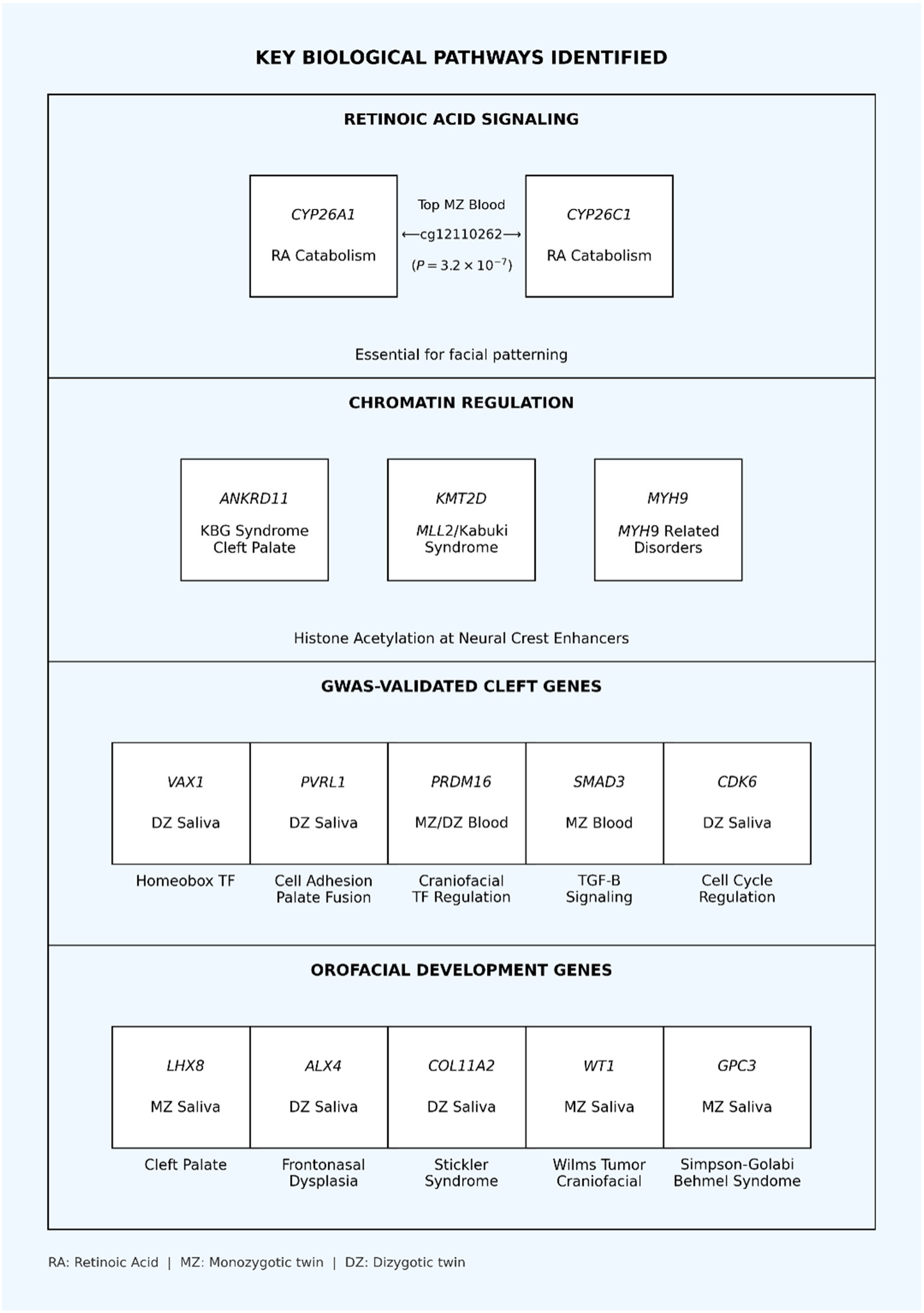
Key biological pathways implicated by differential methylation findings. Summary schematic illustrating the major biological pathways and gene categories identified through pathway enrichment analysis of top DMCs.

Biological pathway analysis identified several relevant functional categories. Retinoic acid signaling was prominently implicated through the *CYP26A1* DMR, with differential methylation at both an enhancer element and the core promoter suggesting direct transcriptional impact on RA catabolism. *TGF*-beta/*BMP* signaling, essential for palatal shelf elevation and fusion, was represented by *SMAD3* and related pathway components. Chromatin regulation emerged as a theme through *ANKRD11 and KMT2D/MLL2* (mutations cause Kabuki syndrome with cleft palate). Neural crest development genes, including multiple homeobox transcription factors (*VAX1, LHX8, ALX4*), were enriched across analyses.

## Discussion

This study represents the largest epigenome-wide association study utilizing both monozygotic and dizygotic twins discordant for nonsyndromic cleft lip with or without cleft palate. Our findings provide several important insights into the epigenetic architecture of this common congenital malformation.

The identification of a differentially methylated region at the *CYP26A1* locus as the top finding in MZ blood provides compelling evidence for retinoic acid signaling in cleft etiology. This DMR comprises two CpG sites with complementary regulatory functions: cg12110262 (P = 3.21 x 10^-7^) lies within an annotated craniofacial enhancer in the intergenic region, while cg15055355 (P = 1.39 x 10^-3^) is positioned just 21 bp upstream of the *CYP26A1* transcription start site, directly within the core promoter. This architecture—differential methylation at both a distal enhancer and the proximal promoter—suggests coordinated epigenetic dysregulation that could directly impact *CYP26A1* transcriptional output.

Retinoic acid is a critical morphogen for craniofacial development, establishing anteroposterior gradients essential for facial patterning during embryogenesis ^20,25^. *CYP26A1* catalyzes RA degradation, creating RA-free zones necessary for proper tissue specification. Both excess and deficiency of RA cause craniofacial malformations in animal models, including cleft palate ^19,26,27^. The finding of differential methylation at two regulatory elements controlling *CYP26A1* expression, identified in genetically identical MZ twins, strongly implicates environmental or stochastic perturbation of this critical pathway. The promoter-proximal location of cg15055355 (21 bp upstream of the TSS) places this CpG within the region where DNA methylation is most directly linked to transcriptional silencing, suggesting a plausible mechanism for altered gene expression.

This finding has potential clinical implications. Epidemiological studies have associated both maternal vitamin A deficiency and excess with cleft risk ^28,29^. Our data suggest that epigenetic dysregulation of RA catabolism may mediate these associations. The coordinated nature of the differential methylation, affecting both enhancer and promoter elements, suggests a mechanism whereby environmental perturbations during critical developmental windows could lead to sustained alterations in RA pathway regulation (Fig. 5). Furthermore, the persistence of methylation differences at this locus in adult blood samples suggests that developmental epigenetic perturbations may leave lasting molecular signatures, potentially serving as biomarkers of developmental exposures.

**Figure 5.**
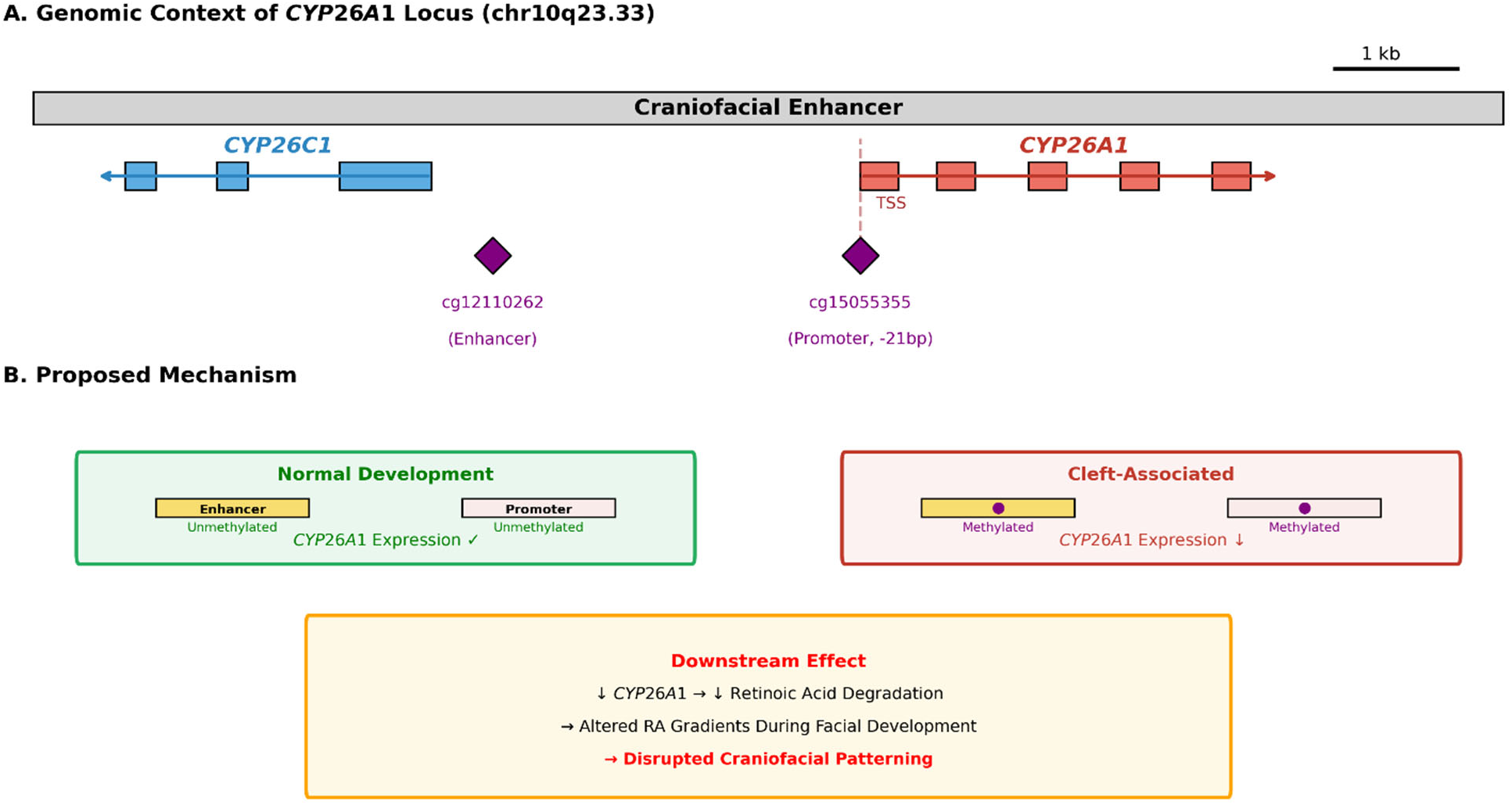
CYP26A1 differentially methylated region: genomic architecture and proposed mechanism. Detailed view of the top finding in MZ blood analysis showing the CYP26A1 differentially methylated region (DMR) on chromosome 10q23.33.

The identification of *ANKRD11* as another top MZ blood hit validates our approach and provides mechanistic insights. *ANKRD11* functions as a chromatin regulator, modulating histone acetylation at gene enhancers essential for neural crest development ^22^. Mutations cause KBG syndrome featuring craniofacial abnormalities and cleft palate in a subset of patients ^21^. The epigenetic dysregulation of *ANKRD11* in genetically identical twins suggests that chromatin regulatory mechanisms may be vulnerable to environmental perturbation, with downstream effects on craniofacial developmental programs.

A key observation is the minimal overlap between MZ and DZ findings, with only 7 CpGs shared across all analyses. This pattern has important implications for understanding cleft etiology. Another observation was that MZ analyses consistently identified more DMCs than DZ analyses across both tissues (Fig. 2a). This pattern is counterintuitive, as one might expect the genetic differences between DZ twins to generate additional methylation variation compared to genetically identical MZ twins. However, this finding is consistent with the concept of variance methylation quantitative trait loci (VmeQTLs) ^17,18^—genetic variants that modulate epigenetic plasticity rather than mean methylation levels. Under this model, MZ twins may be detecting CpG sites with high environmental sensitivity, where genetically identical individuals can nonetheless diverge in methylation due to differential exposures or developmental stochasticity. In contrast, genetic variation in DZ twins may drive methylation toward genotype-specific levels at meQTL loci, potentially masking environmental variation and reducing the detectable signal. The greater DMC yield in MZ twins thus suggests that purely environmental or stochastic epigenetic variation is substantial and detectable when genetic background is controlled. This epigenetic heterogeneity has implications for prevention strategies, as different individuals may require different interventions based on their genetic background.

The striking enrichment of DMCs in craniofacial enhancers, particularly in MZ blood (37.2%), suggests that environmental epigenetic effects may preferentially target tissue-specific regulatory elements rather than promoters or gene bodies. Enhancers are increasingly recognized as key mediators of developmental gene regulation and disease susceptibility ^30–32^. The *CYP26A1* DMR is notable in spanning both an enhancer element and the core promoter, suggesting that in some cases, environmental epigenetic perturbations may coordinately affect multiple regulatory elements controlling the same gene. The lower enhancer enrichment in saliva, despite higher overall DMC counts, may reflect the different cellular composition and developmental origins of these tissues.

The validation of our approach through identification of known cleft susceptibility genes provides confidence in the biological relevance of findings. Multiple GWAS-validated genes including *VAX1, PVRL1, PRDM16*, and *SMAD3* were identified, along with syndromic cleft genes such as *ANKRD11* and *ALX4*. This convergence of EWAS findings with established genetic associations suggests shared biological pathways underlying both genetic and epigenetic contributions to cleft susceptibility.

Several limitations warrant consideration. DNA methylation was measured in blood and saliva samples collected postnatally, after cleft development was complete. While some methylation differences may represent causal factors preserved in somatic tissues, others may reflect consequences of the malformation or its treatment. Blood and saliva are necessarily proxies for embryonic facial tissues, which cannot be directly sampled in humans. However, growing evidence supports the presence of systemic epigenetic alterations associated with developmental outcomes ^33^, and our enrichment in craniofacial enhancers suggests biological relevance. Functional validation studies, such as reporter assays examining the effect of methylation at the *CYP26A1* promoter on transcriptional activity, would strengthen the mechanistic interpretation of our findings.

The modest sample size, while substantial for a discordant twin study, limits statistical power to detect small effect sizes. The effect sizes observed (typically 1-5% methylation differences) are consistent with other EWAS of complex traits ^34^. Future studies with larger cohorts and functional validation will be important for confirming and extending these findings. Replication in independent cleft cohorts, potentially including non-twin samples, would strengthen the evidence for identified associations.

In conclusion, our EWAS of discordant MZ and DZ twins provides novel evidence for epigenetic contributions to nonsyndromic cleft lip/palate. The identification of a differentially methylated region at the *CYP26A1* locus, with CpG sites at both a craniofacial enhancer and the core promoter (21 bp upstream of the TSS), strongly implicates retinoic acid catabolism in disease etiology and suggests a direct mechanism for transcriptional dysregulation. The *ANKRD11* finding connects epigenetic variation to chromatin regulatory mechanisms. The distinct patterns between MZ and DZ twins reveal epigenetic heterogeneity in cleft etiology with potential implications for personalized prevention strategies. These findings advance our understanding of how environmental factors may contribute to this common birth defect through epigenetic mechanisms.

## Methods

### Study Participants and Sample Collection

This study utilized DNA samples extracted from 32 pairs of monozygotic (blood = 24; saliva=8) and 22 pairs of dizygotic twins (blood = 9; saliva = 13) discordant for NSCL/P (Table 1). In addition to the 54 pairs of discordant, we included 8 pairs of monozygotic twins that were concordant for NSCL/P (both twins affected in each pair) as controls. All samples have been previously collected as part of different studies, all approved by their respective IRBs and by the University of Iowa IRB (IRB#201906713). Inclusion criteria included: (1) confirmed twin status by clinical assessment or genetic testing, (2) one twin affected with nonsyndromic cleft lip with or without cleft palate, (3) co-twin unaffected by orofacial clefts, and (4) availability of both blood and saliva samples. Exclusion criteria included syndromic forms of clefting, other major congenital anomalies, or insufficient sample quality for methylation analysis. The participants’ age at the time of sample collection varied across pairs; however, the co-twin design, in which each twin was paired with their respective co-twin, automatically controls for age difference between each case and their paired control (within-pair comparisons). Twin zygosity was confirmed using genetic analysis of 59 polymorphic markers included in the Illumina EPIC array. Monozygotic status was confirmed by concordance at >99% of tested genetic markers, while dizygotic twins showed the expected 50% concordance pattern.

### Sample Quality

DNA quality was assessed with DropSense96™ and quantified with the Qubit™ dsDNA High Sensitivity Range Assay Kit (ThermoFisher Scientific). For each specimen, 500 ng of genomic DNA was bisulfite converted with the EZ DNA Methylation™ Kit (Zymo Research) according to manufacturer’s protocol.

### DNA Methylation Analysis

Genome-wide DNA methylation profiles of 32 MZ twin pairs clinically discordant for NSCL/P were generated using Illumina’s Infinium Methylation EPIC BeadChip assay (EPIC array version 1) (Illumina, San Diego, CA, USA). The assay determines DNA methylation levels in more than 850,000 CpG sites and provides coverage of CpG islands, RefSeq genes, ENCODE open chromatin, ENCODE transcription factor-binding sites, and FANTOM5 enhancers. The assay was performed according to the manufacturer’s instructions and scanned with the Illumina iScan System. To avoid batch effects, both members of a selected twin pair were assayed on the same array and inter-batch duplicate samples were used as internal controls. As expected, duplicate samples showed high degrees of correlation of beta values (r^2^>0.99).

### Initial Processing and Quality Control

All downstream analysis steps were performed in R (version 4.3.1). Initial processing, removal of low-quality targets, and generation of beta values were accomplished using the openSesame workflow (sesame package version 1.18.4) with default parameters ^35^.

Because beta values exhibit heteroscedasticity (unequal variance), beta values were transformed into M-values using the BetaValueToMValue function. M-values were used in all statistical tests described below. Before proceeding to subsequent analyses, the distribution of beta-values and M-values were visually confirmed using histograms generated with ggplot2 (version 3.4.3) and quantile-quantile (Q-Q) plots generated with the stat_qq function.

To correct for differences in cell composition among samples, EpiDISH (version 2.16.0) was used ^36^. For blood samples, the cent12CT.m reference was used, which contains data on 12 blood cell subtypes. For saliva samples, the centEpiFibIC.m reference was used, which contains data on epithelial cells, fibroblasts, and total immune cells.

### Data Visualization and Clustering

We performed Principal Component Analysis (PCA) using the matrix of M-values as input to the prcomp function in R (stats package version 4.3.1) to visualize similarity among samples. The first principal components were visualized in two dimensions with ggplot2 (version 3.4.3). A scree plot was used to assess the variance explained by each principal component.

For selected experimental groups with many samples, all principal components were fed into a Uniform Manifold Approximation and Projection (UMAP) algorithm (package version 0.2.10.0) ^37^, which generated a two-dimensional representation of the samples. UMAP is a non-linear dimensional reduction technique that typically outperforms PCA for large sample size data.

### Statistical Analysis

To test for statistical differences in methylation status between affected and unaffected individuals, the limma package (version 3.56.2) was used to create linear models ^38^ containing the following variables: sample group (affected or unaffected), sex, demographics, age, and twin pair as a blocking factor to account for the paired nature of the data. To test for the potential of hidden batch effects, we used the SmartSVA package (version 0.1.3) ^39^. For sample groups exhibiting hidden batch effects, surrogate variables (SVs) computed using SmartSVA were added to the linear model before performing statistical analysis with limma. To confirm statistical results generated using limma, paired t-tests were performed for some experimental comparisons using the matrixStats package (version 1.0.0). In all cases, histograms of p-values were generated with ggplot2 to visually confirm that the p-value distribution followed the expected pattern.

Statistical significance was assessed using empirical Bayes moderated t-statistics, with false discovery rate (FDR) correction for multiple testing using the Benjamini-Hochberg method. To further prioritize the list of differentially methylated sites, we used a percentile-based approach, filtering our list to include only those methylation sites in which both the raw p-value and the absolute value of the difference in M-values between affected and unaffected were in the top 1% of the distribution.

To reduce the contribution of genotype-driven methylation differences to the MZ discordant blood analysis, CpG sites exhibiting differential methylation in concordant MZ twin pairs were used as a filter. The eight concordant MZ twin pairs (both twins affected) were analyzed in parallel using the same limma linear model pipeline described above, applied to blood samples (due to sample availability). CpG sites reaching nominal significance (P < 0.05) in the concordant MZ blood analysis were excluded from the final list of differentially methylated CpGs in the MZ discordant blood analysis. Because both twins in a concordant pair are affected, methylation differences identified in this group are unlikely to reflect disease-associated epigenetic changes and more likely represent technical, stochastic, or genotype-driven variation unrelated to discordance. This filtering step is intended to enrich the MZ discordant blood results for CpG sites reflecting environmental or developmental epigenetic contributions to NSCL/P.

### Comparison of Different Experimental Groups

To compare the resulting CpG sites generated in various statistical analyses, we first generated lists of CpG identifiers and visualized overlaps with Venn diagrams using the VennDiagram package (version 1.7.3). The statistical significance (hypergeometric p-value) of observed set overlaps was assessed using the SuperExactTest package (version 1.1.0) ^40^. Two additional approaches were used to identify commonalities between different CpG lists. First, downstream and upstream target genes were assigned to each CpG using information from the GREAT database (accessed with the rGREAT package, version 1.99) ^41^. We then looked for overlaps in target genes among different CpG lists. Second, after assigning each CpG to a genomic bin (size 10,000 bp), we counted the number of CpGs in each bin and looked for overlaps between experimental groups.

### Functional Annotation and Pathway Analysis

To map differentially methylated CpG sites to chromosomal locations and other genomic features, we used the chip manifest file (version v-1-0-b5) from Illumina. To facilitate sorting and joining to other annotation databases (e.g., ClinVar, OMIM), data were converted into GRanges objects using the GenomicRanges package (version 1.52.0), and the plyranges package (version 1.20.0) was used to perform common range operations such as joins and counting overlaps ^42,43^. When necessary, genomic coordinates were converted between hg19 and hg38 builds using the liftOver executable from UCSC ^44^. To determine if differentially methylated CpGs located in intergenic regions might be associated with relevant enhancer sequences, we mapped the distance between each CpG and a curated list of 4,346 known craniofacial-associated superenhancers ^31^. To assess if CpGs are located near genes with craniofacial expression patterns, we downloaded expression data from FaceBase (accession FB00001358, processed data count files) ^45^. Gene ontology and pathway enrichment analyses were performed using DAVID and Reactome databases ^46,47^.

## Data Availability

All data produced will be made available online and are currently available upon reasonable request to the authors.

## Acknowledgments

This work was supported by the National Institute of Dental and Craniofacial Research, Eunice Kennedy Shriver National Institute of Child Health and Human Development, (grant number NIDCR K01DE027995, R37DE08559, U01DE020057, R01DE012472, R21DE016930, R01DE014667, R01DE028300).

## Authors Contributions

ALP, HLK, JCM, MLM, XJX, and LMMU contributed to the study conception and design. Sample collection, material preparation, data collection, and analysis were performed by ALP, LD, HLK, XJX, EZ, AB, AW, MLM, JCM and LMMU. ALP is the corresponding author and principal investigator of the study. The first draft of the manuscript was written by ALP, HLK, and LMMU and all remaining authors reviewed and commented on previous versions of the manuscript. All authors read and approved of the final manuscript.

## Ethical Statement

All samples were obtained in accordance with prior study protocols, following their respective approval by the University of Iowa and local Institutional Review Boards (IRBs), and with informed consent provided by participants, parents or guardians.

